# Day-to-day Variability Indices improve utility of Oscillometry in Paediatric Asthma

**DOI:** 10.1101/2023.11.05.23298080

**Authors:** Jane Hoang, Alexander Wong, Kate Hardaker, Sashritha Peiris, Ediane de Queiroz Andrade, Anneliese Blaxland, Penny Field, Dominic Fitzgerald, Geshani Jayasuriya, Chetan Pandit, Hiran Selvadurai, Greg King, Cindy Thamrin, Paul D Robinson

## Abstract

**Background:** Oscillometry may be a feasible and sensitive tool for objective remote monitoring of paediatric asthma.

**Methods:** School-aged cohorts of healthy, well controlled and poorly controlled asthma (defined as ≥2 exacerbations within the preceding 12m) performed daily home-based oscillometry for 3-4 months (C-100 tremoflo, Thorasys Ltd), alongside objective measures of asthma control (ACQ weekly, ACT monthly), medication use (Hailie®) and exacerbations.

Day-to-day variability was calculated as coefficient of variation (CV) for resistance at 5Hz (R5), reactance (X5) and Area under reactance curve (AX). We examined the ability to differentiate asthma from health and correlations with asthma control and exacerbation burden. Clinical exacerbation phenotypes were examined using principal component analysis and k-means clustering of oscillometry, symptoms, breathing parameters and symptoms.

**Results:** Feasibility was 74.9 ± 16.0% in health (n=13, over 93.7 ± 16.2 days) and 80.6 ± 12.9% in asthma (n=42, over 101.6 ± 24.9 days; 17 well controlled 27 poor asthma control). Significantly higher day-to-day variability in all oscillometry indices occurred in asthma, vs. health, and with worsening asthma control. CV R5 when clinically stable (CV R5 stable) was the best discriminator of asthma from health (AUC 0.87, p=0.00001). CV R5 correlated with all measures of asthma control and asthma exacerbation burden, r 0.41-0.52 (all p<0.01). Two exacerbation phenotypes were found based on oscillometry data in the pre-exacerbation period, characterised by severity of impairment of R5, X5, AX and CV R5 (n=12 more severe). Findings were similar using post-exacerbation period oscillometry data (n=8 more severe). Symptoms did not differ across clusters.

**Conclusions:** Home-based oscillometry monitoring was highly feasible over extended periods in school-aged asthmatics. Utility was evidenced by improved ability to differentiate asthma from health, reflect asthma control and exacerbation burden and phenotype exacerbations.

**TAKE HOME MESSAGES:**

- It is highly feasible to perform daily parent-supervised FOT monitoring for extended periods up to 4 months duration in school-aged children
- In contrast to single-session based oscillometry indices, day-to-day variability in oscillometry indices were significantly higher in children with asthma compared to healthy controls, and differentiated levels of asthma control. The best performing parameter was CV R5.
- All day-to-day variability indices correlated with measures of asthma control, with the best performing parameter CV R5 during stable periods (i.e., not including exacerbation periods).
- Amongst asthmatics, day-to-day variability was greater during exacerbation periods than during non-exacerbation periods. Day-to-day variability correlated with measures of exacerbation burden, with the strongest correlations observed with CV R5 during stable periods
- Day-to-day variability identified two distinct clusters of exacerbation, which were not identified by conventional measures or symptom based assessment.

**AUTHOR CONTRIBUTIONS:**

- Conception and design: PDR, CT, GGK
- Recruitment, acquisition, analysis and/or interpretation of data: JH, AW, KH, SP, EdQA, AB, PF, DF, GJ, CP, CT, GGK, PDR
- Writing the manuscript or revising it critically: JH, AW, KH, SP, EdQA, AB, PF, DF, GJ, CP, HS, GGK, CT, PDR

## Introduction

The disease burden of asthma for society is significant. Asthma affects 10% of children and adults (1, 2) with huge annual healthcare costs (>$24billion in Australia, >$80billion in the US) (3). Despite recent advances in treatment, asthma remains a significant cause of morbidity and mortality: 50% of children with asthma have inadequately controlled symptoms(4); it accounts for almost 3% of paediatric emergency department visits and hospitalisations annually in the US (3); is the 16^th^ highest cause of global years lived with disability, and ranked in the top 20 in childhood disability-adjusted life years (5), with the greatest burden between 10-14 years (1). Asthma deaths remain a significant concern because many are preventable with the strongest risk factors being poor asthma control and ongoing exacerbations(6, 7). Conventional symptom-based guideline approaches to obtaining good asthma control continue to be inadequate, limited by factors such as poor symptom perception or reporting by children and parents(8). Furthermore, it is increasingly recognized that asthma exacerbations can be highly heterogeneous, and better phenotyping may facilitate better targeted, more effective treatment(9). In the 2022 GINA report, it recognized the rapid increasing use of digital technology, telemedicine and telehealthcare but highlighted that greater research is required to outline how these can be beneficial(10).

Asthma is characterised by chronic inflammation of the airways and *variable* airway obstruction. Yet current objective lung function-based tools, such as peak expiratory flow (PEF) or spirometry have limited utility, especially in children. It is difficult to obtain consistent technically acceptable data for FEV1 and PEF in children due to the effort-dependent nature of the test. The demonstrated value, in adult data, of looking at *variability* of PEF to predict exacerbations or loss of control(11) does not translate to children. Written daily PEF diaries are unreliable in children(12), and PEF variability (measured using electronic PEF meters) correlates poorly with asthma severity(13). In addition, there is limited sensitivity to detect this variable airflow obstruction: FEV1 lies within the normal range in the majority of asthmatics and may remain so during acute exacerbation(14); In contrast, oscillometry is an effort-independent tidal breathing test which addresses these issues. It has strong feasibility in children down to preschool age(15) and enhanced sensitivity, in comparison to FEV1, to detect bronchodilator response(16, 17) and airway hyper-responsiveness(18, 19). We have previously shown that in an asthma camp setting, under physician-supervision, measurement of day-to-day variability in oscillometry indices differentiated asthmatics from healthy controls, differentiated between levels of asthma severity and asthma control(20). FEV_1_ and PEF failed to do this within that cohort.

There are currently no studies in children outlining feasibility and utility of daily monitoring with oscillometry, under parental supervision, in the home setting. In this study, the primary aim was to determine feasibility of measurement in this setting over a prolonged period of up to four months, whilst secondary aims were to determine whether day-to-day variability in oscillometry-derived respiratory system resistance and reactance at 5Hz (R5 and X5, respectively) (i) differentiates asthma from healthy controls, (ii) correlates with validated measures of asthma control and exacerbation frequency, and finally (iii) to explore the clinical utility in phenotyping asthma exacerbations. We hypothesised that oscillometry-based home monitoring would be highly feasible, that day-to-day variability in oscillometry indices would be higher in asthmatics, and in those with worse asthma control and higher exacerbation frequency and would provide ability to phenotype exacerbations over and above conventional measures including symptoms.

## Methods

### Subjects and Study Design

The subjects presented in this manuscript were recruited across two studies, both with ethics approval from the Sydney Children’s Hospital Network (SCHN) Human Research Ethics Committee (14/SCHN/572 and 2019/ETH13753). Written informed consent/assent was obtained from all parents/guardians and children. The children were all aged 8-18 years, and recruited between April 2016 and December 2022. Asthmatics all had physician-diagnosed asthma managed within a tertiary paediatric chest and asthma clinic at The Children’s Hospital at Westmead (CHW), New South Wales. Asthmatics within the first study were enrolled in an observational study and underwent home monitoring for 3-4 months, with preferential recruitment for those with a history of “poorly controlled asthma” (defined as a history of reported exacerbations in the preceding 12 months). The second study enrolled subjects with a treating physician’s assessment of “well controlled asthma” based on good reported asthma control for ≥6 months. To define normal day-to-day variability, healthy control subjects, defined as those with no known history of preterm birth, chronic respiratory condition or previous admission to hospital for respiratory illness or regular respiratory medication use were recruited for a two month period of home monitoring.

### Baseline assessment

A detailed assessment of baseline lung function was performed at study entry, consisting of Multiple and Single Breath Washout (MBW and SBW, respectively), oscillometry and spirometry in all subjects. Nitrogen (N2) based MBW and SBW (Exhalyzer D, Spiroware v3.1.6, ECO MEDICS AG, Dürnten, Switzerland, reanalysed in v3.3.1) were performed in triplicate according to ERS/ATS consensus inert gas washout statement(21). Outcomes reported were LCI, FRC, S_cond_ and S_acin_. Oscillometry training and baseline assessment was performed as triplicate 60s recordings using a pseudorandom waveform containing 5-37Hz (tremoFlo® C-100 Airwave Oscillometry System™, THORASYS® Thoracic Medical Systems Inc, Montreal, Canada, v1.04 build 43) according to current ERS technical standards at the time(22, 23). Outcomes reported were airway resistance and reactance at 5Hz (R5 and X5, respectively) and Area under reactance curve (AX) as z scores using recently published paediatric reference data(24). Spirometry(25) was performed using JAEGER® (Vyntus Masterscreen PNEUMO, IOS module, Germany, SentrySuite software version 3.0) and outcomes (FEV1, FEV_1_/FVC and FEF_25-75_) were reported as z-scores using GLI reference equations(26). Questionnaires performed were the Asthma Control Questionnaire (ACQ) (27), Asthma Control Test (ACT) and the Paediatric Asthma Quality of Life Questionnaire (PAQLQ).

### Home monitoring period

Following oscillometry training and device setup in the subjects’ home, families were instructed to perform triplicate measurements once a day (at a time convenient for the family) for ≥3-4 months. Where suitable electronic medication data loggers existed for the medications used (SmartInhalers™, Adherium Ltd., Auckland, New Zealand) these were installed onto both preventer and reliever asthma medications. Subjects completed an electronic daily questionnaire immediately prior to measurement and were contacted weekly by videoconferencing to review oscillometry technique and SmartInhaler™ technique proficiency, perform an ACQ and record details of any exacerbations. Asthma exacerbations were defined according to Virchow et al(28). ACT and PAQLQ were recorded monthly. Oscillometry data were reviewed by research staff at the completion of the monitoring period. Technically acceptable test occasions were defined as those with three recordings each containing at least five breaths that were free of technical artefact, consistent with guideline recommendations(22). Breaths containing artefacts were excluded. Only days with technically acceptable data were included in analyses.

### Data analyses

Home monitoring feasibility was characterised as the percentage of days with technically acceptable measurements, expressed as the percentage of the total monitoring period, and percentage of the total monitoring period excluding days where the family/child were away from the home where equipment had been installed. Day-to-day variability of oscillometry indices was calculated as coefficient of variance (CV%) of data from the entire monitoring period. To avoid the contribution of lung function behaviour during exacerbations, “stable” day-to-day variability indices were also calculated by excluding oscillometry data collected during defined exacerbation periods (e.g. “R5 CV stable”). Based on the distribution of data, differences between groups were compared using t-tests or Mann Whitney U, or when more than two groups were compared by ANOVA or Kruskal-Wallis tests, respectively.

Correlations were assessed using Pearson or Spearman correlation. Receiver-operator characteristic (ROC) curves were used to evaluate the performance of day-to-day variability in oscillometry indices to differentiate health vs. disease. This was compared to the performance of baseline oscillometry parameters to differentiate health from disease.

We explored exacerbation phenotypes based on oscillometry indices (including mean and CV%), breathing parameters, symptoms, and medication adherence data in 7-day time windows in the weeks surrounding each exacerbation. Principal component analysis (PCA) followed by k-means clustering were performed to determine the presence of any data-driven clusters. Input variables and patient characteristics were then compared across clusters. The effects of varying number of clusters, window size and timing were also examined in sensitivity analyses. Further details of the methods used in the study are provided in the online supplement (OLS).

## Results

### Study Population

Thirteen healthy and 42 asthmatic subjects participated, their baseline characteristics and lung function are shown in Table 1. Baseline demographics were well matched between health and asthma, as were several asthma characteristics amongst asthmatics. Almost all asthmatic subjects were sensitized on skin prick testing to aeroallergens, with the majority having other diagnosed atopic disease. Amongst poorly controlled asthmatics, most were at step 4 or higher of current GINA treatment guidelines. Poorly controlled asthmatics had higher numbers of exacerbations in the preceding year, and at the time of the baseline assessment, evidence of worse asthma control as evidenced by lower ACT and higher ACQ scores. Asthmatics had lower lung function at enrolment than healthy controls (Table 2). Unexpectedly worse lung function was observed in the well-controlled group compared to those with a history of increased exacerbations – across all spirometry outcomes, when expressed as z scores (all p<0.05), as well as MBW variable Scond*VT (p=0.002).

**Table 1.** Baseline Demographics and Asthma Characteristics of the cohort.

|  |  |  | Healthy | Asthmatics |  |  |
| --- | --- | --- | --- | --- | --- | --- |
|  |  |  |  | All asthmatics | Well controlled | Poorly controlled |
| Demographic |  |  |  |  |  |  |
|  | Number of subjects (Male, %) |  | 13 (39%) | 42 (64%) | 17 (64%) | 25 (64%) |
|  | Age at study entry (years) |  | 12.3 ± 2.4 | 13.11 ± 2.52 | 13.21 ± 2.56 | 13.05 ± 2.56 |
|  | Height z-score |  | 0.75 ± 1.12 | 0.04 ± 0.87 | 0.12 ± 0.89 | -0.01 ± 0.87 |
|  | Weight z-score |  | 0.54 ± 1.12 | 0.39 ± 0.97 | 0.31 ± 0.88 | 0.44 ± 1.05 |
|  | BMI |  | 0.75 ± 1.12 | 0.04 ± 0.87 | 0.12 ± 0.89 | -0.01 ± 0.87 |
|  |  | Overweight (85.0 – 94.9 Centile) | 0 | 11 (26) | 4 (24) | 7 (28) |
|  |  | Obesity (≥95.0 Centile) | 2 (15) | 5 (12) | 1 (6) | 4 (16) |
| Asthma characteristics |  |  |  |  |  |  |
|  | Age of Asthma Diagnosis (years) (n=38) |  | - | 3.29 ± 1.99 | 3.49 ± 1.74 | 3.15 ± 2.16 |
|  | Asthma Duration (years) |  | - | 9.81 ± 3.24 | 9.69 ± 2.85 | 9.89 ± 3.54 |
|  | Atopy (Positive skin prick test) |  | 0 | 38 (90) | 15 (88) | 23 (92) |
|  | Allergy Rhinitis (n=39) |  | 0 | 32 (76) | 15 (88) | 17 (68) |
|  | Eczema (n=39) |  | 0 | 25 (60) | 11 (65) | 14 (56) |
|  | Persistent Asthma (n=38) |  | 0 | 41 (98) | 17 (100) | 24 (96) |
| Asthma exacerbations |  |  |  |  |  |  |
|  | Total Exacerbations in last 12 months |  | - | 4.05 ± 4.45 | 0.64 ± 1.15 | 5.96 ± 4.47*** |
|  | OCS Courses in last 12 month |  | - | 3.15 ± 4.13- | 0.64 ± 0.93 | 4.56 ± 4.57*** |
|  | Hospital Admissions in last 12 months |  | - | 1.69 ± 3.17- | 0.29 ± 0.61 | 2.48 ± 3.73** |
| Asthma medication |  |  |  |  |  |  |
|  | GINA Stage |  |  |  |  |  |
|  |  | Stage 3 | 0 | 13 (31) | 8(47) | 5 (20)** |
|  |  | Stage 4 | 0 | 15 (36) | 4 (24) | 11 (44)** |
|  |  | Stage 5 | 0 | 7 (16) | 1(6) | 6 (24)** |
| Questionnaires |  |  |  |  |  |  |
|  | Asthma Control Questionnaire (ACQ) |  | 0.00 ± 0.00 | 0.75 ± 0.82 | 0.37 ± 0.34 | 0.97 ± 0.94*** |
|  | Asthma Control Test (ACT) |  | 25.00 ± 0.00 | 19.7 ± 4.18 | 20.44 ± 3.81 | 19.21 ± 4.41*** |

Data shown as number (%) or Mean ± SD. All variables were compared across the 3 groups (Healthy control, Well-controlled, and poorly controlled) using ANOVA or Fisher exact test where appropriated, \*p<0.05, \*\*p<0.01, \*\*\*p<0.001, \*\*\*\*p<0.0001.

**Table 2.** Baseline Lung function of the cohort.

|  |  | Healthy | Asthmatics |  |  |
| --- | --- | --- | --- | --- | --- |
|  |  |  | All asthmatics | Well controlled | Poorly controlled |
| Spirometry |  |  |  |  |  |
|  | FEV<sub>1</sub> % Pred | 98.5 ± 8.6 | 92.4 ± 17.1 | 87.7 ± 19.4 | 95.6 ± 15.0 |
|  | FVC % Pred | 100.0 ± 11.6 | 102.0 ± 14.9 | 98.2 ± 14.9 | 104.6 ± 14.6 |
|  | FEV<sub>1</sub>/FVC | 86.4 ± 7.0 | 78.7 ± 9.5 | 76.8 ± 9.9 | 80 ± 9.2* |
|  | FEF<sub>25-75</sub> % Pred | 94.3 ± 14.7 | 73.4 ± 27.6 | 67.1 ± 28.7 | 77.7 ± 26.6* |
| Spirometry |  |  |  |  |  |
|  | FEV<sub>1</sub> z-score | -0.12 ± 0.74 | -0.69 ± 1.45 | -1.41 ± 1.63 | -0.37 ± 1.27* |
|  | FEV<sub>1</sub>/FVC z-score | -0.01 ± 1.00 | -1.29 ± 1.23 | -2.03 ± 0.93 | -1.03 ± 1.21* |
|  | FEF<sub>25-75</sub> z-score | -0.27 ± 0.69 | -1.43 ± 1.44 | -2.24 ± 1.34 | -1.07 ± 1.35* |
| Multiple Breath Washout |  |  |  |  |  |
|  | LCI (turnovers) | 6.21 ± 0.30 | 7.28 ± 1.64 | 7.56 ± 2.00 | 7.09 ± 1.35 |
|  | Scond*VT | 0.026 ± 0.011 | 0.051 ± 0.022 | 0.054 ± 0.024 | 0.048 ± 0.020** |
|  | Sacin*VT | 0.061 ± 0.020 | 0.078 ± 0.054 | 0.079 ± 0.042 | 0.077 ± 0.062 |
| Single Breath Washout |  |  |  |  |  |
|  | SIII | 1.31 ± 0.50 | 1.59 ± 0.93 | 1.62 ± 0.92 | 1.56 ± 0.96 |
|  | SIII * VC | 3.69 ± 1.05 | 4.48 ± 3.07 | 4.42 ± 2.18 | 4.52 ± 3.61 |
|  | CV | 0.235 ± 0.168 | 0.268 ± 0.236 | 0.200 ± 0.145 | 0.315 ± 0.276 |
| Oscillometry (n=42) |  |  |  |  |  |
|  | R5 z-score | -0.15 ± 1.78 | -0.06 ± 1.41 | 0.42 ± 1.86 | -0.40 ± 0.9 |
|  | X5 z-score | 0.11 ± 1.87 | -0.03 ± 1.56 | -0.56 ± 2.06 | 0.33 ± 1.00 |
|  | AX z-score | -0.99 ± 3.09 | 1.05 ± 5.33 | 2.91 ± 7.51 | -0.22 ± 2.60 |

Data shown as Mean ± SD. % Pred; percent predicted. All variables were compared across the 3 groups (Healthy control, Well-controlled, and poorly controlled) using ANOVA or Fisher exact test where appropriated, \*p≤0.05, \*\*p≤0.01, \*\*\*p≤0.001, \*\*\*\*p≤0.0001.

### Feasibility of Home monitoring

Overall, 55 monitoring periods were completed by 13 healthy subjects and 42 asthmatic subjects (17 with well controlled asthma control and 27 with poor asthma control). Strong feasibility was observed across the groups (Table 3) based on both percentage of total days enrolled or as percentage of days not away from home. Feasibility results were similar between the groups.

**Table 3:**
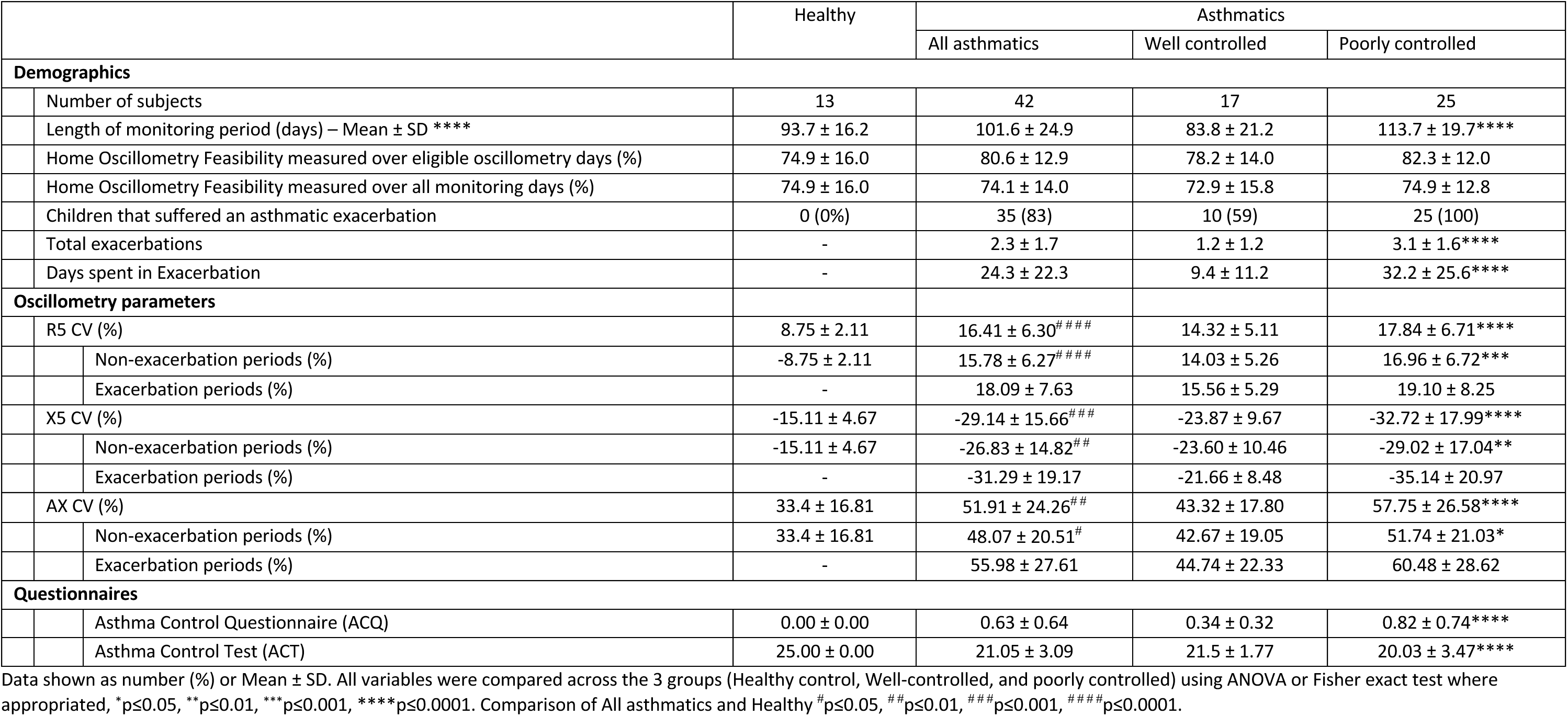
Oscillometry Home monitoring data.

### Comparing health vs asthma

Significantly higher day-to-day variability in all oscillometry indices were observed in asthmatic subjects, compared to healthy subjects (Table 3). In contrast baseline oscillometry indices’ z-scores were not different (Table 2). Across the three groups of healthy, well controlled and poorly controlled asthmatics a progressive increase in day-to-day variability was observed, with significant differences across the groups in all indices regardless of whether data was from the entire monitoring period or only from stable periods (Figure 1). The strongest statistically significant difference was observed with R5 CV (all p<0.0005), followed by X5 CV (p<0.005) and AX CV (all p<0.05).

**Figure 1.**
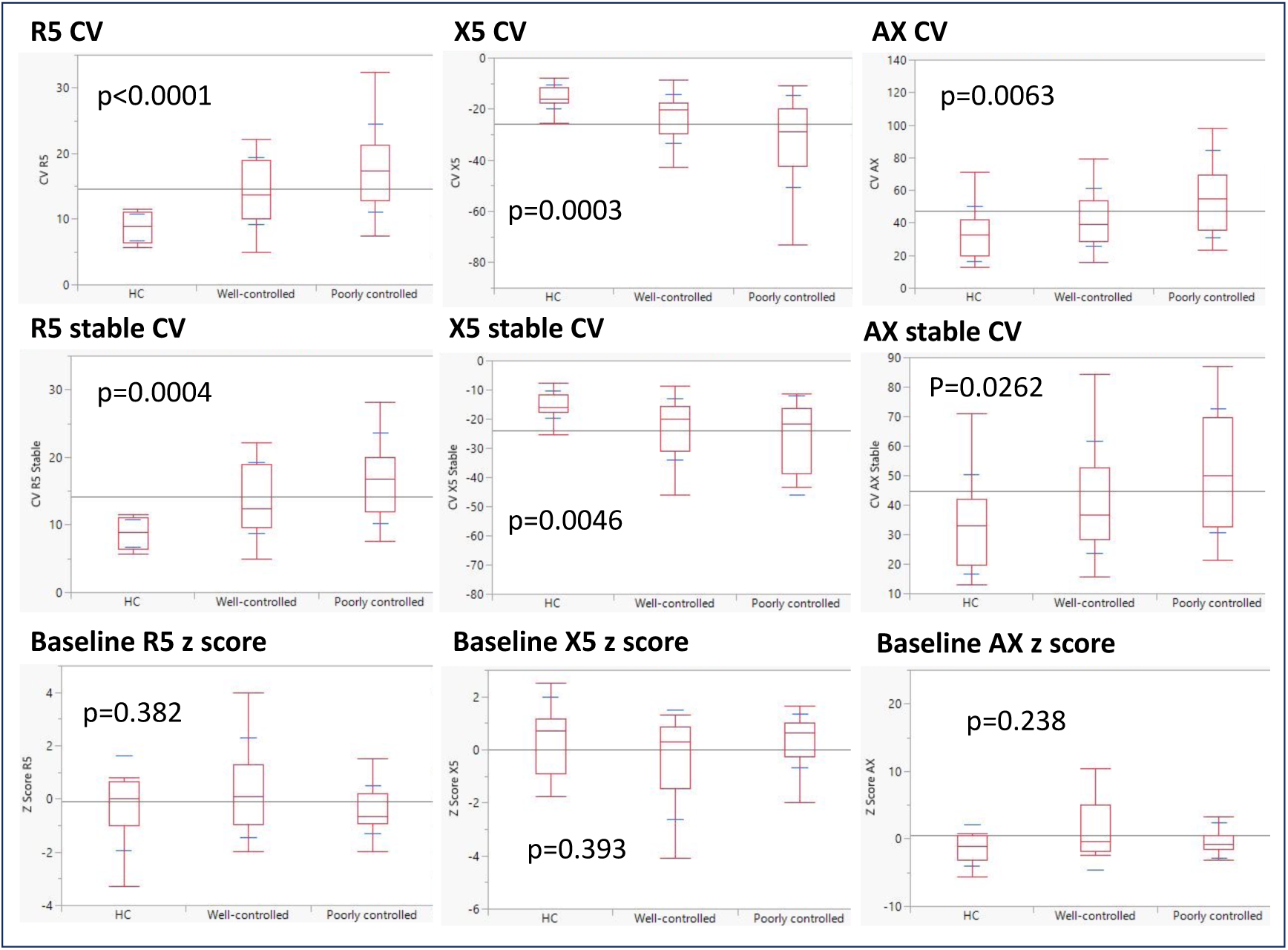
Comparison of day-to-day variability in oscillometry indices and baseline oscillometry indices across healthy, well controlled and poorly controlled asthma subjects. Footnote: Box and whisker plots display median, first and third quartiles and range for each parameter. CV, coefficient of variation; HC, heathy controls

ROC analysis showed high predictive values (defined as AUC ≥0.80) (Figure 2) for CV R5 and CV X5 and CV R5 stable, with the highest AUC observed for CV R5 stable (AUC 0.87).

**Figure 2.**
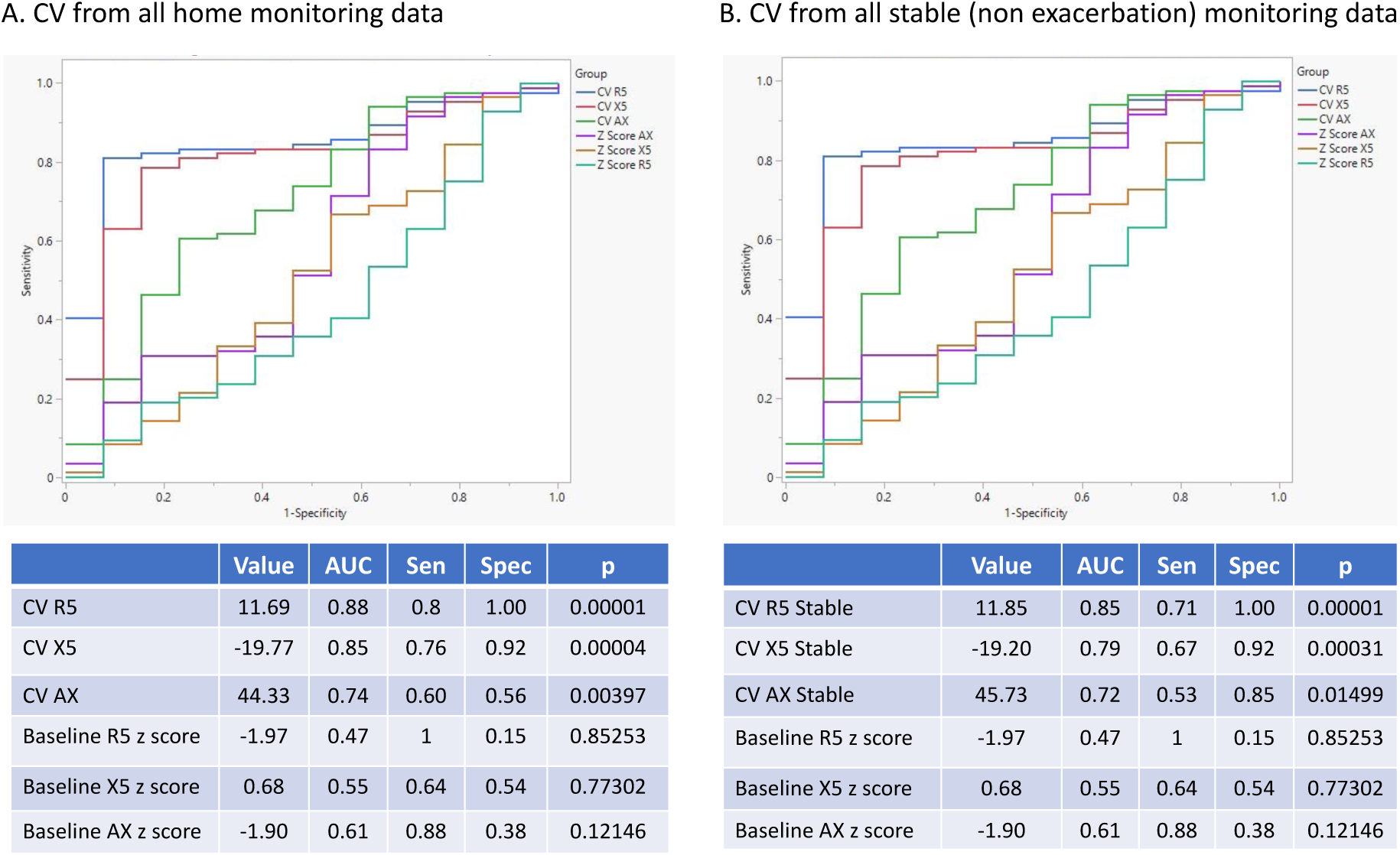
Comparison of ability of day-to-day variability in oscillometry indices to differentiate asthma from healthy subjects, in comparison to baseline oscillometry indices. Footnote: CV, coefficient of variation; AUC, area under the curve; Sen, sensitivity; Spec, specificity.

Sensitivity and specificity were 0.80 and 1.00, respectively, for CV R5. The AUC of baseline oscillometry indices were between 0.47-0.61 and were not statistically significant.

Objective measures of asthma control during the monitoring period (mean ACQ and mean ACT) also differed between well-controlled and poorly controlled asthmatic subjects (Table 2). All day-to-day variability oscillometry indices correlated with measures of asthma control (Table 4, Figure 3). The strongest correlations, achieving statistical significance across all measures of asthma control, were observed with CV R5 (r0.41-0.51, all p<0.01). AX-based indices also correlated with both ACT and ACQ but showed weaker correlations (0.32-0.38, p<0.05), whilst X5-based indices correlated significantly only with ACQ. Similar patterns were observed for correlations of questionnaire reported asthma control outside of exacerbation periods, further strengthening correlations observed when compared to “stable” period day-to-day variability indices.

**Figure 3.**
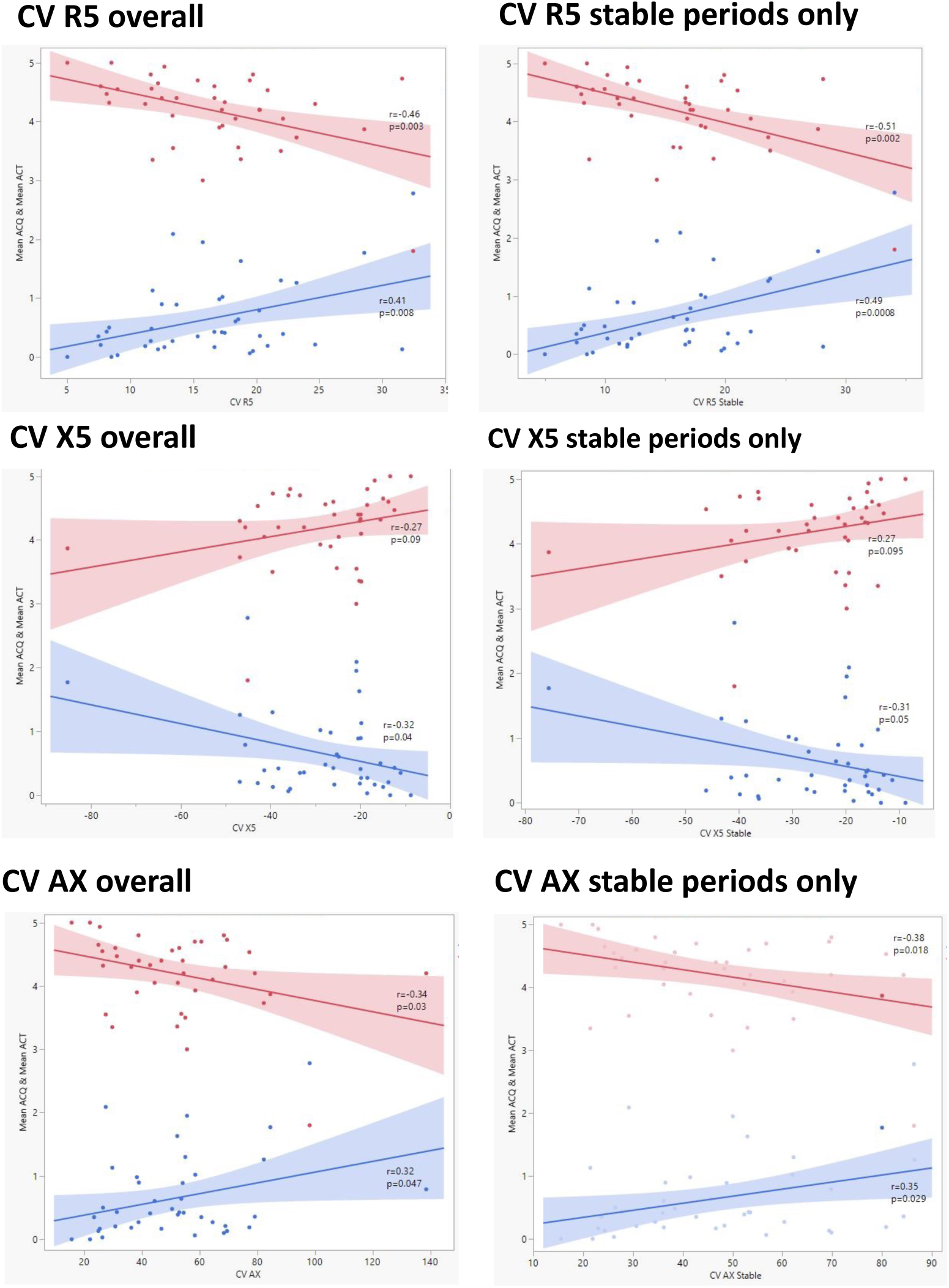
Correlation analysis of day-to-day variability in Oscillometry indices with asthma control as assessed by ACQ (red) and ACT (blue). Footnote: Shaded areas represent the 95% confidence intervals of the correlation. ACQ and ACT expressed as a mean value across the monitoring period for each participant.

**Table 4.** Correlation analysis for day-to-day variability parameters and measures of asthma control and exacerbation.

|  | Mean ACQ | Mean ACQ Stable | Mean ACT | Mean ACT Stable | Exac Days | Exac number |
| --- | --- | --- | --- | --- | --- | --- |
| CV R5 | 0.41** | 0.45** | -0.46** | -0.42** | 0.49** | 0.48** |
| CV R5 Stable | 0.49*** | 0.50*** | -0.51*** | -0.46** | 0.52*** | 0.49*** |
| CV X5 | -0.32* | -0.44** | 0.27 | 0.27 | -0.44** | -0.49** |
| CV X5 Stable | -0.31* | -0.43** | 0.27 | 0.29 | -0.40** | -0.42** |
| CV AX | 0.32* | 0.30 | -0.34* | -0.40* | 0.35* | 0.42** |
| CV AX Stable | 0.35* | 0.35* | -0.38* | -0.51** | 0.38* | 0.37* |

\*p<0.05, \*\*p<0.01, \*\*\*p<0.001, \*\*\*\*p<0.0001; Exac, exacerbation.

Within the monitoring period, the exacerbation burden differed between the groups (p<0.0001), with the largest values occurring in poorly controlled subjects, in whom exacerbation number was mean±SD 3.1±1.6 and percent days spent in exacerbation was 32.2±25.6% of days (Table 3). Day-to-day variability was greater for all oscillometry indices during exacerbation periods, compared to non-exacerbation periods. All day-to-day variability indices correlated with measures of exacerbation burden, both expressed as exacerbation number or percent days spent in exacerbation (Table 4). The strongest correlations were again observed with CV R5 stable, reflecting oscillometry behaviour during non-exacerbation monitoring periods.

### Exacerbation phenotyping

When considering the pre-exacerbation period, the optimum window for clustering (i.e. yielding the highest silhouette coefficient of 0.513, see OLS) began 10 days prior to exacerbation onset. Two clusters of exacerbations were identified on the basis of principal component 1 (Table E1, Figure E1), with a smaller cluster (Cluster 2, n = 12) typified by higher R5, X5 and AX and higher R5 and X5 day-to-day variabilities compared to the larger cluster (Cluster 1, n=53), but no differences in symptom scores (Table E2). Cluster 2 was associated with participants who had more abnormal S_cond_, and more participants used LABA and LTRA at study entry (Table E3). A similar pattern was observed for the post-exacerbation onset period, where the optimum window began 2 days after exacerbation onset (silhouette coefficient: 0.593). Again, two clusters were identified on the basis of principal component 1 (Table E4, Figure E2), whereby a smaller cluster (Cluster 2, n=8) had higher R5, X5, AX and also day-to-day variability across all these indices compared to the larger cluster (Cluster 1, n=65), but no differences in symptoms scores (Table E5). Here also, Cluster 2 was associated with participants with poorer S_cond_, and more of them used LTRA at study entry (Table E6). Note the difference in number of exacerbations between the pre- and post-exacerbation onset periods is due to different windows excluded on the basis of missing data points.

Importantly in all cases, including symptoms, controller adherence or breathing data in the cluster analyses decreased the silhouette coefficient, suggesting oscillometry was the most significant classifier of exacerbation phenotypes.

## Discussion

This study provides evidence that daily parent-supervised oscillometry monitoring is not only highly feasible in school-aged children in the home setting for prolonged periods but that it provides additional insight into important aspects of asthma missed by conventional hospital-based testing. Day-to-day variability in oscillometry indices was increased in asthma, compared to healthy controls, and amongst asthmatics was greater in those with poor asthma control. It enabled oscillometry to effectively differentiate asthma from health, in contrast to single-session hospital-based measurements. It showed more consistent correlation with validated measures of asthma control and exacerbation burden. Day-to-day variability in R5, assessed during periods of non-exacerbation, performed best amongst these oscillometry indices. Finally, oscillometry indices of day-to-day variability identified two distinct asthma exacerbation phenotypes, in contrast to conventional approaches including symptoms, revealing a small cluster of patients with worse day-to-day variability (CV R5 and CV X5), as well as higher day-to-day variability during exacerbation. This may indicate a more severe asthma exacerbations phenotype.

Strong feasibility of remote monitoring, in homes up to 700km away from the tertiary hospital, allowed us to confirm two important findings observed in our initial pilot study conducted on site within an asthma camp setting(29), but now performed over a much longer period of monitoring, with a commercial oscillometry device, under parental supervision. Day-to-day variability in oscillometry indices was higher in those with asthma, compared to healthy controls. Additional ROC-based analyses outlined the strong ability (i.e., AUC values >0.8) to differentiate asthma from health, in a larger cohort than our original study. Furthermore, amongst these asthmatics day-to-day variability correlated with validated questionnaire-based assessments of asthma control. A key component of asthma control, and an established risk factor for poor asthma outcomes, is the occurrence of asthma exacerbations – in this study the use of daily electronic diaries, electronic medication monitoring and weekly remote follow ups, gave us strong resolution to record exacerbations. Day-to-day variability correlated with two measures of asthma exacerbation burden – number of asthma exacerbations and proportion of days spent in exacerbation.

Importantly these relationships with asthma control and exacerbation burden persisted when examining non-exacerbation day-to-day variability, suggesting the same level of insight is being provided when examining periods separate from the exacerbations themselves. Future work will determine the ability of oscillometry to provide an early signal for exacerbation in children, but is suggested by the pattern of change observed within subjects in these cohorts(30).

Given that the objective measures of asthma control used in this study are determined by symptoms, and therefore symptom perception, the moderate strength (as opposed to strong) correlations observed within our data suggest that day-to-day variability in oscillometry contains additional information not captured by symptoms alone. This is further supported by the additional value of oscillometry in phenotyping exacerbations within our dataset. The investigation of ability to phenotype exacerbations better than conventional approaches was explored in response to an urgent call in a NEJM editorial to better understand asthma exacerbations and not treat them as a single entity(9). Our exploratory work provided evidence for distinct exacerbation phenotypes, with the main contributor to this phenotyping being oscillometry data, with symptoms adding relatively little value.

Regardless of timing surrounding the exacerbation, we found a small cluster of exacerbations with higher oscillometry indices and higher day-to-day variability in oscillometry. These tended to occur in participants who had worse ventilation heterogeneity at baseline, perhaps suggesting an underlying physiological mechanism distinguishing exacerbation phenotypes. This represents an important initial first step in identifying different types of exacerbations, with oscillometry appearing to provide a more objective assessment than subjective symptom scores. Take together these results represent an important first step towards developing strategies which help mitigate the issues of variation in patient perception and reporting of symptoms.

Strengths of this study include its prospective longitudinal cohort study design and the steps taken to ensure good quality data for oscillometry, medication logging, and asthma control and identification of exacerbations. By targeting children with a recent history of exacerbations in one of the cohorts, we were able to capture a large number of exacerbations during the subsequent home monitoring period. The length of the monitoring period ensured adequate oscillometry data and identification of start and end points of exacerbation to allow accurate calculation of variability for both exacerbation and non-exacerbation periods. By including a range of asthma control, in comparison to a contemporaneous control cohort, we provide insight into the potential utility across the spectrum of asthma and provide a strong basis for further studies in this area.

In conclusion, the results of this study outline the strong feasibility of extended periods of home-based oscillometry monitoring in school-aged asthmatics, and its additional utility, compared to conventional lung function assessment. It improved the ability of oscillometry to differentiate asthma from health, to reflect asthma control, and amongst asthmatics those with both a preceding risk, and concurrent risk of asthma exacerbation. Future studies will need to confirm whether oscillometry day-to-day variability provides an early signal of loss of asthma control and whether management decisions based on these day-to-day fluctuations during both non-exacerbation and exacerbation periods can improve important patient reported outcome measures such as asthma control, exacerbation burden and exacerbation severity, to reduce the enormous health cost of asthma.

## Supporting information

Online supplement Paediatric asthma home oscillometry monitoring

## Data Availability

All data produced in the present study are available upon reasonable request to the authors

## Acknowledgements

Research study funding provided by the Sydney Medical School Foundation, the Ross Trust, and Asthma Australia. Thorasys Ltd. (the manufacturer of the commercial oscillometry device used in this study) provided oscillometry devices and technical support in kind for the duration of the study, and Adherium Ltd. (the manufacturer of the electronic compliance monitoring device for the inhaled asthma medication used in this study) provided technical support in kind for the duration of the study. The Lloyd Nuttall Asthma Research Fellowship for funding protected research time for the senior investigator (PR). John Reynolds for his assistance in establishing remote backup of data.

