## Supplementary material for "Day-to-day Variability Indices improve utility of Oscillometry in Paediatric Asthma": Online supplement Paediatric asthma home oscillometry monitoring

### 1. Lung function techniques

#### *1.1 Multiple breath washout (MBW)*

Technical acceptability of MBW trials was assessed using published quality control criteria(1), developed based on inert gas ERS/ATS inert gas washout consensus statement recommendations(2). $Sn_{III}$ analysis involved visual inspection and estimation of phase III slopes ($S_{III}$) for each breath, with $S_{cond}$ and $S_{acin}$ calculated per trial and averaged across the three acceptable trials. Only trials with acceptable $S_{III}$ for first breaths and at least 2/3 of breaths between 1.5-6.0 lung turnovers were accepted, where the value of 1 lung turnover was equivalent to the subjects' FRC for that trial. Reference values to define abnormality for LCI, FRC and $Sn_{III}$ indices ($S_{cond}$ and $S_{acin}$) used were equipment and software version specific(3). Due to the recent description of a cross-talk error between the oxygen and carbon dioxide sensors in this equipment, which affects the outcomes describe in this manuscript and has been corrected in updated software (v3.3.1), all MBW and SBW data was re-analysed in the updated software version(4, 5). Published upper limits of normal offs for LCI, $S_{cond}$ and $S_{acin}$ (6) were used, updated to Spiroware v3.3.1.

#### *1.2 Single breath washout (SBW)*

Technical acceptability of SBW trials was assessed using published quality control criteria, developed based on inert gas ERS/ATS inert gas washout consensus statement recommendations(2). Visual inspection and estimation of phase III slopes ($S_{III}$) and closing volume for each trial was performed.

Technically acceptable test occasions were defined as those with three trials with vital capacity (VC) measurements within 10% of the highest VC value across the SBW tests. Mean $S_{III}$ (%/L) and $S_{III}$ x expiratory VC (%) across the three acceptable trials were reported.

2. **Measures of asthma control**

ACQ was developed and validated in adults (7), was later validated for use in children aged 6-16 years (8). The questionnaire has a recall period of one week and consists of seven questions, asking for breathlessness, nocturnal awakenings, symptoms when waking up, activity limitation, wheeze, frequency of rescue medication, and pre-bronchodilator $FEV_1$% predicted. The items are scored from 0 (optimal) to 6 (worst) and are equally weighted. The ACQ score is calculated as the mean of the seven items and ranges from 0 (well controlled) to 6 (extremely poorly controlled*)*. In this study we used a shortened version, where $FEV_1$ was omitted, which has been shown to perform as well as the complete version(9). Asthma control was graded using ACQ < 0.75 as indicating well controlled whilst ACQ ≥ 1.5 indicating poorly controlled(10).

The ACT was developed for children and adults 12 years and older and has a recall time of four weeks(11). It consists of five questions on shortness of breath, awakenings at night, limitation of activity, rescue SABA use, and patient rating of asthma control. Each item is scored on a scale of 1–5 and the scores are added, to a total score of 5–25. The C-ACT has been developed for children aged 4-11 years(12). It is a seven-item questionnaire that divided into two parts. The first part filled in by the child using a visual scale from 0-3 across four questions on perception of asthma control, limitations of activities, coughing, and nocturnal awakenings. The second part is three questions scored from 0-5 by the parent/caregiver on daytime symptoms, daytime wheezing, and awakenings at night. The sum of the C-ACT score is the sum of all scores (0-27). Asthma control is graded using recognised thresholds (13): controlled asthma (ACT≥20 points) and uncontrolled asthma (ACT≤19 points) (11, 12, 14).

### 3. **Asthma Exacerbations**

Exacerbation records were used to determine start and end of exacerbations according to the an implementable definition described by Virchow et al(15)  based on earlier published ATS/ERS recommendations (16). An exacerbation was defined as ≥1 of the following criteria:

- Nocturnal awakening(s) due to asthma requiring SABA for 2 consecutive nights or increase from baseline in daily symptom score on 2 consecutive days
- Increase from baseline in occasions of SABA use on 2 consecutive days (minimum increase: 4 puffs/day)
- Visit to the emergency room/trial site for asthma treatment not requiring systemic corticosteroids

A severe exacerbation was defined as a deterioration in asthma symptoms requiring the use of systemic corticosteroids, ED presentation or hospitalisation. A minimum of 7 days between exacerbations was required to differentiate separate exacerbations and should a moderate exacerbation progress into a severe exacerbation, the exacerbation was considered an escalation of symptoms and was classified as one severe exacerbation. Should a period of severe exacerbation require more than one distinct course of rescue corticosteroids and tapering-off period, each course required was considered a separate severe exacerbation. Transient deterioration in symptoms or transient SABA use (≤1 day), that did not result in an emergency department visit were not recorded as exacerbations. Prophylactic SABA use before exercise was accounted for and only rescue SABA use in response to asthma symptoms was deemed relevant to determining an exacerbation.

### 4. **Phenotyping asthma exacerbations**

#### *4.1 Identification of exacerbations and calculation of time windows*

The onset of each exacerbation was defined as the first day on which either increase in asthma symptoms, increase in rescue medication, or nocturnal awakening occurred for moderate exacerbations, or the first day of increase in symptoms leading to oral steroids or ED/hospitalization for severe exacerbations.

For each patient, we then examined the variations in oscillometry, breathing parameters, symptoms and adherence surrounding the start of exacerbation, using 7-day overlapping time windows progressed one day at a time. For each 7-day window, we calculated the following: mean R5, X5 and AX; their coefficients of variance (CV), inspiratory and expiratory R5 and X5, tidal volume (VT), respiratory rate (RR), ACQ5 and controller adherence.

Days with missing data were replaced with data from the previous day (last observation carried forward), upto a maximum of 2 days prior. However, windows were excluded if ≥50% of data was missing. This was the basis for choosing a window size of 7 days, as this allowed CV to still be quantified with <50% valid data. This also meant that total number of exacerbations included in different analyses may have varied slightly due to excluded time windows.

*5.2 Data reduction and classification*

Principal component analysis (PCA) was used to reduce the dimensionality of the large number of variables of interest (17). The outcome of PCA is a new set of variables (principal components), which are linear, uncorrelated combinations of the original variables, which were ranked in decreasing order of their contribution to the data variance. The goal is to explain the variance in the data in as few variables as possible.

K-Means clustering was then applied to the first 3 components arising out of PCA, to identify and classify clusters of points within the data (possible phenotypes) (18, 19). For a fixed number of clusters (decided a priori), this approach determines the optimal clusters by using a repetitive algorithm to maximise the distance between clusters, which can be visualized in 3D.

The silhouette coefficient (SC)(20) was used as a metric for performance of the cluster analysis. It is a measure of how similar any given data point is to other points within its own cluster, compared to data points in other clusters.

#### *5.3 Sensitivity analyses*

We examined the effect of varying window sizes (from 2 to 14 days) and timing (from 21 days prior ranging to 14 days after exacerbation onset) on the main results. We also examined the effect of number of clusters, and of including/excluding additional variables. The SC was used to determine the optimum performing results.

#### *5.4 Results*

In the pre-exacerbation period, the optimum 7-day window for clustering (SC=0.513) began 10 days prior to exacerbation onset, with only including the oscillometry indices R5, X5, AX, R5 CV, X5 CV and AX CV, and with number of clusters = 2. The contributions from these variables to the top 3 principal components are shown in Table E1 – in the final results, clusters were split on component 1 alone.

**Table E1.** Contribution of oscillometry indices to principal components obtained from the optimum pre-exacerbation window.

| Variable | Component 1 | Component 2 | Component 3 |
|---|---|---|---|
| R5 | 0.174 | 0.1614 | 0.0273 |
| X5 | 0.1822 | 0.1534 | 0.0203 |
| AX | 0.1958 | 0.1301 | 0.006 |
| CV R5 | 0.1546 | 0.176 | 0.4737 |
| CV X5 | 0.1686 | 0.1601 | 0.3771 |
| CV AX | 0.1247 | 0.2189 | 0.0956 |

The identified clusters are displayed in Figure E1. Between-cluster comparisons of oscillometry, breathing parameters, and symptoms are provided in Table E2, whereas differences in patient characteristics at study entry are provided in Table E3.

**Figure E1.** Clusters identified on 3 component PCA from the optimum pre-exacerbation window.

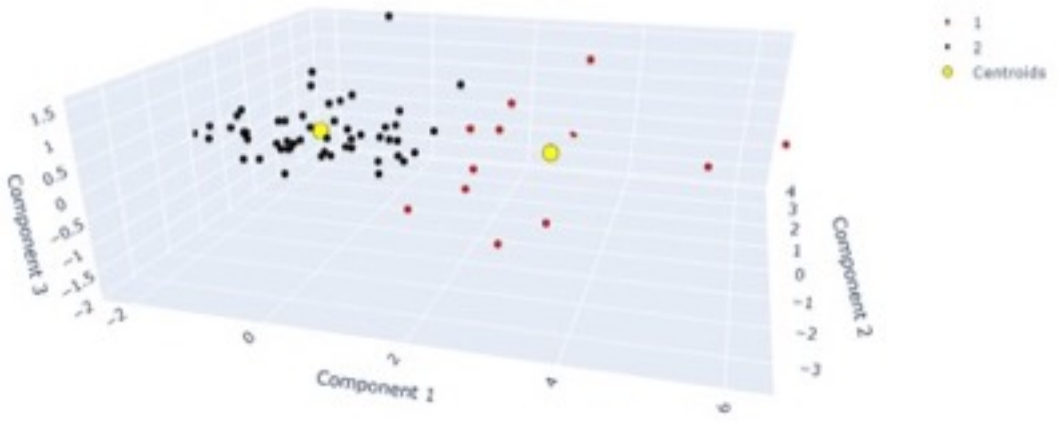

**Table E2.** Differences in oscillometry, breathing parameters and symptoms between the two clusters based on the optimal pre-exacerbation window.

| | **Cluster 1** (n=53) | **Cluster 2** (n=12) | **P value** |
|---|---|---|---|
| R5, $cmH_2O\ s\ L^{-1}$ | 4.18 (0.82) | 6.75 (1.36) | **<0.001** |
| X5, $cmH_2O\ s\ L^{-1}$ | -1.47 (0.48) | -2.99 (0.79) | **<0.001** |
| AX, $cmH_2O\ L^{-1}$ | 9.8 (5.8) | 31.6 (9.9) | **<0.001** |
| CV R5, % | 11.4 (5.2) | 20.2 (11.5) | **0.024** |
| CV X5, % | -15.4 (8.7) | -36.1 (17.1) | **0.001** |
| CV AX, % | 33.8 (16.7) | 51.0 (27.6) | 0.059 |
| Trouble breathing | 0.378 (0.459) | 0.646 (0.821) | 0.295 |
| Asthma bother | 0.326 (0.408) | 0.520 (0.659) | 0.346 |
| Activities limited | 0.207 (0.313) | 0.302 (0.347) | 0.397 |
| VT, L | 0.704 (0.225) | 0.671 (0.339) | 0.753 |
| RR, breaths per minute | 21.9 (6.0) | 23.1 (4.6) | 0.421 |

Footnote: Mean(SD) and P value for paired t-test shown unless otherwise indicated.

**Table E3.** Differences in patient baseline and overall characteristics between the two clusters based on the optimal pre-exacerbation window.

| | **Cluster 1** (n=53) | **Cluster 2** (n=12) | **P value** |
|---|---|---|---|
| Age at study entry, years | 13.1 (2.6) | 11.9 (2.3) | 0.124 |
| Gender, M:F | 30:23 | 11:1 | **0.023†** |
| BMI, kg $m^{-2}$ | 21.1 (3.8) | 20.8 (4.3) | 0.814 |
| GINA stage | 4.0 (0.8) | 3.8 (0.8) | 0.480 |
| $S_{cond}$, Z-score | 3.76 (2.10) | 5.33 (1.08) | **0.001** |
| $S_{acin}$, Z-score | 0.35 (1.7) | -0.22 (2.71) | 0.502 |
| ACQ5 | 0.9 (0.8) | 1.5 (1.0) | 0.070 |
| Mean adherence, % | 81.6 (10.3) | 86.9 (9.9) | 0.118 |
| LABA use, yes:no | 29:24 | 11:1 | **0.018†** |
| LTRA use, yes:no | 21:32 | 10:2 | **0.006†** |
| Exacerbations/month | 0.3 (0.3) | 0.5 (0.4) | 0.068 |

Footnote: Mean(SD) and P value for paired t-test shown unless otherwise indicated. †Chi-square test; n=number of exacerbations; GINA stage – asthma control as defined by global initiative for asthma criteria; LABA – long-acting beta2-agonist; LTRA – leukotriene receptor antagonist.

In the period post exacerbation onset, the optimum 7-day window for clustering (SC=0.598) began 2 days after exacerbation onset, with only including the oscillometry indices R5, X5, AX, R5 CV, X5 CV and AX CV, and with number of clusters = 2. The contributions from these variables to the top 3 principal components are shown in Table E4 – in the final results, clusters were again split on component 1 alone.

**Table E4.** Contribution of oscillometry indices to principal components obtained from the optimum post-exacerbation window.

| Variable | Component 1 | Component 2 | Component 3 |
|---|---|---|---|
| R5 | 0.1713 | 0.1596 | 0.2323 |
| X5 | 0.1657 | 0.1754 | 0.1203 |
| AX | 0.1873 | 0.1386 | 0.0856 |
| CV R5 | 0.1744 | 0.1496 | 0.1005 |
| CV X5 | 0.1636 | 0.1704 | 0.2817 |
| CV AX | 0.1377 | 0.2064 | 0.1797 |

The identified clusters are displayed in Figure E2. Between-cluster comparisons of oscillometry, breathing parameters, and symptoms are provided in Table E5, whereas differences in patient characteristics at study entry are provided in Table E6.

**Figure E2.** Clusters identified on 3 component PCA from the optimum post-exacerbation window

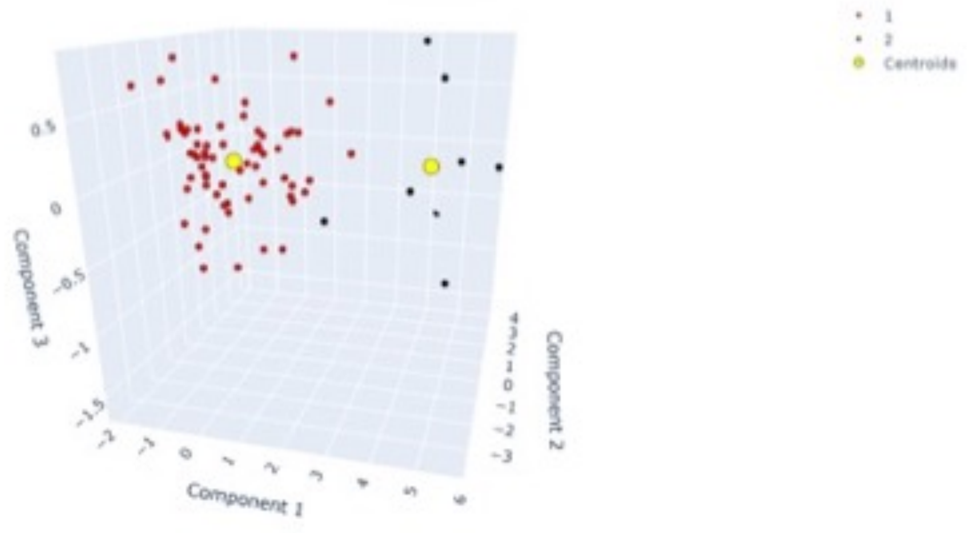

**Table E5.** Differences in oscillometry, breathing parameters and symptoms between the two clusters based on the optimal post-exacerbation window.

| | **Cluster 1** (n=65) | **Cluster 2** (n=8) | **P value** |
|---|---|---|---|
| R5, $cmH_2O\ s\ L^{-1}$ | 4.22 (0.93) | 6.74 (1.32) | **0.001** |
| X5, $cmH_2O\ s\ L^{-1}$ | -1.55 (0.54) | -2.97 (0.80) | **0.001** |
| AX, $cmH_2O\ L^{-1}$ | 10.9 (6.6) | 32.9 (10.0) | **<0.001** |
| CV R5, % | 10.3 (4.7) | 26.8 (9.7) | **0.002** |
| CV X5,% | -15.2 (8.5) | -43.2 (16.5) | **0.002** |
| CV AX,% | 31.3 (16.6) | 70.3 (31.3) | **0.009** |
| Trouble breathing | 0.442 (0.581) | 0.557 (0.839) | 0.716 |
| Asthma bother | 0.428 (0.61) | 0.449 (0.693) | 0.937 |
| Activities limited | 0.307 (0.479) | 0.357 (0.484) | 0.789 |
| VT, L | 0.697 (0.23) | 0.659 (0.225) | 0.664 |
| RR, breaths per minute | 21.9 (5.4) | 21.9 (3.4) | 0.975 |

Footnote: Mean(SD) and P value for paired t-test shown unless otherwise indicated.

**Table E6.** Differences in patient baseline and overall characteristics between the two clusters based on the optimal post-exacerbation window.

| | **Cluster 1** (n=65) | **Cluster 2** (n=8) | **P value** |
|---|---|---|---|
| Age at study entry, years | 12.9 (2.6) | 12.1 (2.2) | 0.350 |
| Gender, M:F | 40:25 | 7:1 | 0.148† |
| BMI, kg $m^{-2}$ | 21.1 (3.9) | 20.8 (4.0) | 0.816 |
| GINA stage | 4.0 (0.8) | 3.9 (0.8) | 0.756 |
| Scond, Z-score | 3.97 (2.02) | 5.45 (1.19) | **0.010** |
| Sacin, Z-score | 0.40 (1.72) | 0.33 (2.94) | 0.948 |
| ACQ5 | 0.9 (0.8) | 1.2 (0.9) | 0.429 |
| Mean adherence, % | 81.8 (10.5) | 86.4 (10.5) | 0.278 |
| LABA use, yes:no | 38:27 | 7:1 | 0.111† |
| LTRA use, yes:no | 30:35 | 7:1 | **0.027**† |
| Exacerbations/month | 0.3 (0.3) | 0.5 (0.4) | 0.262 |

Footnote: Mean(SD) and P value for paired t-test shown unless otherwise indicated. †Chi-square test; n=number of exacerbations; GINA stage – asthma control as defined by global initiative for asthma criteria; LABA – long-acting beta2-agonist; LTRA – leukotriene receptor antagonist.

In general, increasing the number of clusters decreased SC. When varying the window size between 3 to 12 days, keeping the number of clusters = 2 and the timing fixed at the day prior to exacerbation onset, the SC ranged from 0.42 to 0.54 peaking at 5 days; keeping the timing fixed at the day post-exacerbation onset, the SC ranged from 0.40 to 0.53 peaking at 8 days.

In all cases, including symptom, controller adherence or breathing data in the cluster analyses decreased SC, suggesting oscillometry was the most significant classifier of phenotypes.

16. Reddel HK, Taylor DR, Bateman ED, Boulet LP, Boushey HA, Busse WW, Casale TB, Chanez P, Enright PL, Gibson PG, de Jongste JC, Kerstjens HA, Lazarus SC, Levy ML, O'Byrne PM, Partridge MR, Pavord ID, Sears MR, Sterk PJ, Stoloff SW, Sullivan SD, Szefler SJ, Thomas MD, Wenzel SE, American Thoracic Society/European Respiratory Society Task Force on Asthma C, Exacerbations. An official American Thoracic Society/European Respiratory Society

statement: asthma control and exacerbations: standardizing endpoints for clinical asthma trials and clinical practice. *Am J Respir Crit Care Med* 2009; 180: 59-99.

17. Wold S, Esbensen K, Geladi P. Principal Component Analysis. *Chemometrics and Intelligent Laboratory Systems* 1987; 2: 37-52.

18. Haldar P, Pavord ID, Shaw DE, Berry MA, Thomas M, Brightling CE, Wardlaw AJ, Green RH. Cluster analysis and clinical asthma phenotypes. *Am J Respir Crit Care Med* 2008; 178: 218-224.

19. Pedregosa F, Varoquaux G, Gramfort A, Michel V, Thirion B, Grisel O, Blondel M, Prettenhofer P, Weiss R, Dubourg V, Vanderplas J, Passos A, Cournapeau D, Brucher M, Perrot M, Duchesnay E. Scikit-learn: Machine Learning in Python. *Journal of Machine Learning Research* 2011; 12: 2285-2830.

20. Kaufman L. Finding groups in data : An introduction to cluster analysis. Hoboken, NJ: Wiley-Interscience; 1990.
